# Real-World Evidence of the Effectiveness and Safety of Generic Tofacitinib in Rheumatoid Arthritis Patients: A Retrospective, Single-Centre Analysis from Western India

**DOI:** 10.1101/2022.04.08.22273582

**Authors:** Sanat Phatak, Aditya Khenat, Mansi Malandkar, Sanjiv Amin

**Author notes:** Correspondence to: Dr Sanat Phatak, KEM Hospital Research Centre, Sardar Moodliar road, Pune 411011.

## Abstract

**Background:** Generic tofacitinib has been available in India for more than a year and is widely used in rheumatoid arthritis (RA) therapy. There is scarce real-world data on its effectiveness and safety from India, especially given infection endemicity.

**Methods:** We retrospectively analysed records (demographic and clinical information, haematology and biochemistry, adverse events) of patients prescribed generic tofacitinib from a single centre in Mumbai, India. Disease activity was calculated using the disease activity score-28 and erythrocyte sedimentation rate (DAS28-ESR) and other tools, and we used paired T-tests for significant response. We defined clinical tofacitinib failure as a composite outcome, including clinician’s decision to change to an alternative disease-modifying anti-rheumatic drug (DMARD) or flare after self-withdrawal. We performed logistic regression and survival analysis for determinants of clinical failure.

**Results:** We reviewed records of 102 patients (92 female; median age: 53 years) with mean RA duration of 146 months. Thirteen had prior treatment with innovator tofacitinib. There was significant improvement in disease activity parameters at a mean duration of 186 days. No serious adverse events were reported; 4 patients had tuberculosis and 19 patients had mild COVID-19 while on treatment. Clinical failure was seen in 25 patients, and mean time to failure on survival analysis was 357 days. No baseline characteristic predicted clinical failure.

**Interpretation:** Generic tofacitinib showed good effectiveness and a tolerable adverse effect profile, despite tuberculosis endemicity andCOVID-19.. Setting up registries would be valuable in gaining more data on generic tofacitinib.

**Key points:**

- There is scarce data from India regarding the use of tofacitinib in rheumatoid arthritis, despite widespread use
- In this retrospective analysis of 102 patients at a single centre, we found tofacitinib monotherapy was efficacious and tolerable.
- Tuberculosis was detected in four and nineteen patients had mild covid.

## Introduction

The therapeutic armamentarium of rheumatoid arthritis (RA) has been enriched considerably with the addition of targeted synthetic-disease-modifying anti-rheumatic drugs (tsDMARDs), including the oral selective Janus kinase (JAK1/JAK3) inhibitor tofacitinib.^(1)^ Multiple large clinical trials and follow-up studies have established the efficacy and safety of tofacitinib in RA. ^(2-5)^ The United States Food and Drug Administration (US-FDA) approved its use for RA in 2012. ^(6)^

RA in Asian regions differs due to different disease prevalence, adverse effect profiles and socio-economic determinants that impact management. India is endemic for tuberculosis (TB), and has seen multiple waves of the recent COVID-19 pandemic.^(7)^ Compared with European ethnic groups Indians have higher metabolic disease susceptibility,^(8)^ which JAK inhibition may worsen.

There are limited data on the use of tofacitinib in India. Indian patients included in the clinical trials of the innovator tofacitinib were found to have similar efficacy but a different infection profile, (primarily TB) in a post-hoc analysis.^(9)^ While randomised controlled trials [RCTs] provide high-quality data that show evidence of causality, they are artificial environments with strict inclusion criteria, close follow-up and monitoring, and fewer comorbidities.^(10)^ Consequently, the interpretability of the results to real-world patients is compromised; a gap bridged by observational studies and registries.^(11)^ Most observational data on tofacitinib comes from the US, where tofacitinib has been available since 2012.

Despite the dearth of information on tofacitinib in Indian patients, paradoxically, there has been an increase in its use recently in India. Though innovator tofacitinib has been available in India since June 2016, generic tofacitinib was introduced iin November 2020, reducing the monthly therapy cost approximately 10-fold. To the best of our knowledge, there is no real-world observational study of the effectiveness and tolerability of generic tofacitinib from India. We conducted a retrospective analysis of patients prescribed generic tofacitinib from a single centre in western India, and report on its efficacy and safety.

## Methods

We reviewed electronic health records of consecutive patients with RA who received tofacitinib, at an independent private rheumatology clinic in Mumbai, India, who were receiving generic tofacitinib. Tofacitinib was given as monotherapy, started mainly due to suboptimal response to previous therapy, economic considerations or convenience of oral medications in comparison with biologic DMARDs (bDMARDs). Active TB (X-ray), hepatitis B (HBsAg and IgM anti-HBc) and hepatitis C infection (anti-HCV) were ruled out prior to starting the medication.

Data collection-Clinical information at the time of presentation as well as the date of initiation of tofacitinib, including comorbidities, anthropometry, haematological and serological were retrieved from electronic records. This was supplemented as necessary by personal interviews or phone calls, especially regarding adverse effects of medications and intercurrent infections.

Clinical assessments: All clinical assessments at each visit were performed by the same clinician. Rheumatoid factor and C-reactive protein (CRP) were measured by immunoturbidimetry, anti-CCP (Anti-cyclic citrullinated peptides) antibodies by the ELISA method, and erythrocyte sedimentation rate (ESR) by the Westergren method.

Outcome measures – For RA disease activity, composite scores, including disease activity score-28 (DAS28), simplified disease activity index (SDAI) and clinical disease activity index (CDAI), were calculated. DAS28 disease activity and the responders were defined as per the European Alliance of Associations for Rheumatology (EULAR) response criteria.

Drug failure- We defined clinical tofacitinib failure as a composite outcome based on the clinician’s decision to change to an alternative DMARD due to nonresponse or disease flare after patient’s self-withdrawal.

### Statistical analysis

Data are presented as frequencies, mean or median, depending upon the type of data. Response to tofacitinib on various parameters was calculated using paired T-tests to detect differences at onset and last follow-up. We then grouped the patients according to low baseline disease activity (EULAR remission or mild disease activity, DAS28 <3.2) and moderate-to-high disease activity (EULAR moderate and high disease activity, DAS28 >3.2) and repeated paired T-tests separately in these subgroups. Logistic regression analyses were performed to evaluate factors associated with tofacitinib clinical failure. For the regression analysis, we included age, body size measurements, duration of disease, baseline disease activity, presence or absence of comorbidities, prior bDMARD exposure, and baseline disease activity as dependent variables. A survival analysis was performed to assess the mean time to clinical failure.

## Ethics

This study received ethics permission from an independent ethics committee who accorded a consent waiver. Confidentiality was maintained using unique identifiers.

## Results

### Patient characteristics

We studied 102 patients (92 females; median age 53 years) (Table 1). Average body mass index (BMI) was 26+7 (37 were overweight, 14 obese). Hypertension was the most common comorbidity (n=29). The mean duration of disease was 146 months and mean DAS28-ESR was 4.79(1.54). Two thirds of patients had fatigue and weight loss. Five had interstitial pneumonia, 4 had ocular sicca, and 1 had rheumatoid vasculitis.

**Table 1:** Baseline characteristics of patients with RA at initiation of generic tofacitinib (n=102)

|  |  |
| --- | --- |
| Gender ratio | Male: 10, Female: 92 |
| Age, years (median, range) | 53 (21–83) |
| Weight, kg (mean, SD) | 61 (13.9) |
| Height, cm (mean, SD) | 155.9 (7.0) |
| *BMI, kg/m<sup>2</sup> (mean, SD) | 26 (7.0) |
| Comorbidities (n) | None: 47<br>Hypertension: 29<br>Type 2 diabetes: 9<br>Hypothyroidism: 12<br>Bronchial asthma: 1<br>Others: 4 |
| RA disease duration, months (median, range) | 146 (9–520) |
| DAS28-ESR (mean, SD) | 4.79 (1.54) |
| EULAR disease activity according to DAS28-ESR (n) | Remission (<2.6): 9<br>Mild (2.6–3.2): 7<br>Moderate (3.2–5.1): 45<br>Severe (>5.1): 41 |
| CDAI (median, range) | 19 (2–60) |
| SDAI (median, range) | 24 (2.4–160) |
| CRP, mg/dL (mean, SD) | 14.3 (22.1) |
| ESR, mm (mean, SD) | 43 (28) |
| Prior treatment (n) | Treatment naïve: 5<br>csDMARDs only: 64<br>csDMARDs+bDMARDs or bDMARDs only: 20<br>csDMARDs+bDMARD+tsDMARD or csDMARDs+tsDMARD: 13 |

\* Body Mass index categories (n) < 18.5 (6); 18.5–24.9 (45); 25–29.9 (37); > 30 (14)csDMARDs (Methotrexate, Leflunomide, Sulfasalazine, Hydroxychloroquine); bDMARDs: (Adalimumab, Infliximab, Tocilizumab, Etanercept, Rituximab); tsDMARD: Tofacitinib (Innovator).

### Tofacitinib treatment

Eighty-four were switched over to generic tofacitinib monotherapy from previous therapy, 13 from innovator tofacitinib while 5 patients were treatment naive (Table 1). First DMARD received was methotrexate (n=81), sulphasalazine (n=16) and leflunomide(3). Seventy four had received two DMARDs. Twenty-nine patients had received at least one bDMARD (20 had TNF α inhibitors, tocilizumab (n=6) and rituximab (n=3)); 12 had received two bDMARDs.

### Treatment with tofacitinib generic

Mean duration of therapy was186 (74–505) days at evaluation. As a group, the patients responded well; significant differences were observed in most disease activity-related parameters. (Table 2). DAS28 responses correlated well with SDAI (r^2^ of 0.815, p=0.000). Forty-seven classified as DAS28 responders. CRP levels, but not ESR, significantly reduced.

**Table 2:** Response to generic tofacitinib

|  | **At onset**<br>**(mean + SD)** | **At**<br>**follow-up**<br>**(mean + SD)** | **Significance**<br>**p-value** |
| --- | --- | --- | --- |
| TJC | 9 ± 7.8 | 5.2 ± 5.7 | 0.000 |
| SJC | 3.4 + 4.3 | 1.3 ± 2.3 | 0.000 |
| DAS28 | 4.79 ± 1.54 | 4.27 ± 1.4 | 0.002 |
| SDAI | 34.4 ± 28.4 | 24.4 ± 20.3 | 0.001 |
| CDAI | 21.4 ± 13.4 | 15.1 ± 11.2 | 0.000 |
| Weight | 61 ± 13.9 | 64 ± 14.0 | 0.000 |
| Laboratory investigations |  |  |  |
| CRP | 14.3 ± 22.1 | 10.6 ± 13.9 | 0.005 |
| ESR | 43± 28 | 42 ± 28 | 0.799 |
| Hb | 11.2 ± 1.47 | 11.2 ± 1.42 | 0.764 |
| WBC | 7004 ±2399 | 7233 ± 2194 | 0.211 |
| Creatinine | 0.7 ± 0.19 | 0.82 ± 0.32 | 0.044 |
| Cholesterol | 174 ± 34 | 192 ± 39.6 | 0.000 |
| SGPT | 21.6 ± 12.1 | 20.8 ± 10.7 | 0.533 |

**Table 3:**
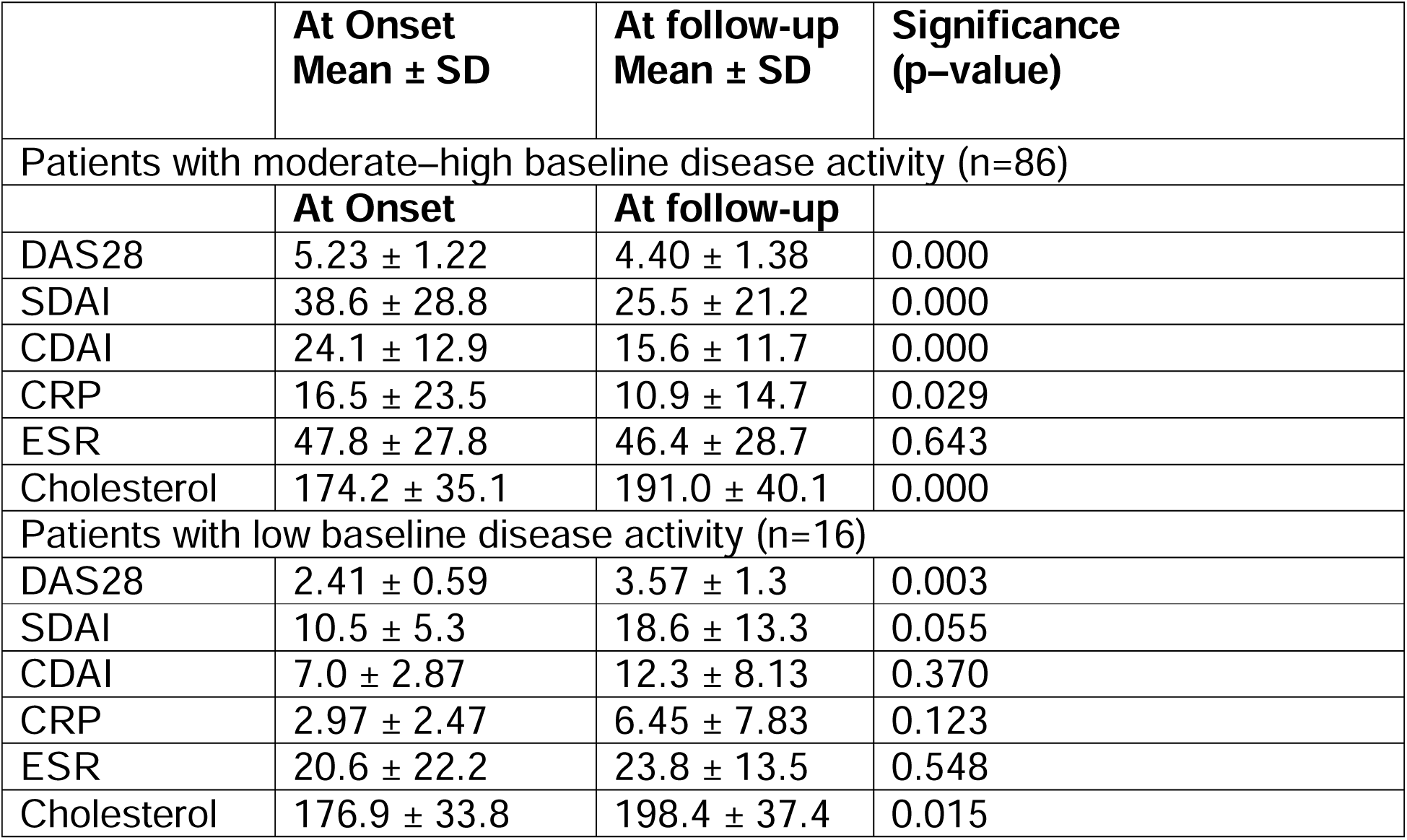
Response to generic tofacitinib in patients segregated by baseline disease activity

These improvements were driven by patients who were switched from other DMARDS (n=89): in these patients, the DAS28-ESR fell from 4.87 to 4.34 (p=0.00), SDAI fell from 34.3 to 25 (p=0.009), and CDAI from 22 to 15 (p=0.007). Patients who were switched to generic tofacitinib from innovator tofacitinib (n=13) did not have a statistically significant change in disease activity parameters (DAS28: 4.2 to 3.8, p=0.328; SDAI 34 to 21; p=0.146) apart from CDAI (16 to 10, p=0.03).

Patients who had moderate-to-high disease activity at baseline (DAS28 >3.2, n=86) improved in all disease-related parameters (Table 2). Disease activity increased (mean DAS28 rise 1.16) in the 16 patients who had low disease activity (DAS28 <3.2) at baseline.

### Adverse events

Fourteen out of 102 patients experienced adverse events (all minor). Most commonly reported AE were GI disorders, paresthesias, rash and itching (Table 4). Two patients had unusual manifestations: claudication and panniculitis. No serious adverse events (thrombosis, cardiovascular events, malignancies, mortality) were reported. Four reported TB; tofacitinib was not stopped in these patients, and anti-TB therapy was given. In 19 patients, a positive SARS-CoV2 nasal RT-PCR was reported while on tofacitinib. All COVID-19 infections were mild, and none required hospitalisation. Tofacitinib was not stopped in any of these patients by the physician. There was no reported case of severe viral infection or herpes zoster in this cohort.

**Table 4:** Safety Summary

| **Adverse events** | **Tofacitinib generic (n=102)** |
| --- | --- |
| **Patients with serious adverse events** | 0 |
| **Patients with adverse events** | 14 (13.75 %) |
| Claudication improved after omission | 1(0.98 %) |
| Right leg panniculitis | 1(0.98 %) |
| Eczema | 1(0.98 %) |
| Giddiness | 1(0.98 %) |
| Rash and itching | 3 (2.94 %) |
| Parasthesia | 3 (2.94 %) |
| Gi disorder | 4 (3.92 %) |
| **Patients discontinuing due to adverse events** | 7 (6.86%) |
| **Discontinue due to pregnancy** | 1 (0.98) |
| **Deaths** | 0 |
| **Adverse events of special interest** |  |
| Serious infections | 0 |
| Herpes zoster | 1 (0.98%) |
| Tuberculosis | 4 (3.92%) |
| MACE | 0 |
| Thrombosis | 0 |
| Malignancy | 0 |
| **Total positive SARS-COV 2 /COVID -19 patients in cohort** | **19 (18.62%)** |

### Metabolic and haematologic adverse effects

As a group, the patients showed increase in weight (mean: 3 kg) and cholesterol (mean: 18 mg/dL) while on tofacitinib. Increase in cholesterol was seen in both low and high disease activity groups at baseline. There was no significant fall in platelet count.

### Tofacitinib failure

Twenty-five had tofacitinib clinical failure: 21 had inadequate response and were switched to a different medication (18 to baricitinib, 2 to adalimumab, and 1 to combination csDMARD), while 4 had flare of RA after self-withdrawal. In addition to these, tofacitinib was stopped in 1 patient because of pregnancy (not included in failure). Patients who failed to respond to generic tofacitinib showed increased DAS28 and SDAI scores (mean increase: 0.50 and 4.3, respectively). Logistic regression analysis revealed no association of failure with characteristics at baseline (age, duration of disease, previous bDMARD use, disease activity at baseline or body phenotype characteristics). The mean time to clinical failure was 357 days (95% CI: 292–439).

## Discussion

This retrospective real-world study analysing Indian patients with RA treated with generic tofacitinib from a single centre in Western India showed a good response to tofacitinib treatment despite long duration, active disease and multiple DMARD failures. There were no serious adverse events during the treatment; 4 patients had TB. COVID-19 was mild in all affected. One-fourth of the patients had a clinically defined failure, which was not predicted by any baseline characteristics at the time of starting tofacitinib therapy.

All our patients received monotherapy; the effectiveness of tofacitinib monotherapy has been reported in the ORAL-START trial. The patients in this study represent a homogenous ethnicity with a wide age range, including 12 patients older than 70 years, unlike the younger Indians in the ORAL studies ^(9)^ but similar the US-based CORRONA registry, in which patients on tofacitinib had a disease duration of >10 years.^(12)^ Additionally, the Indian patients in the clinical trials were bDMARD naïve; our patients therefore represent a more severe disease cohort like is seen in real-world studies. ^(13,14)^

We found that generic tofacitinib led to reduced RA disease activity, also seen in other retrospective studies from various ethnic groups, including those from Japan^(13)^ and Europe^(15)^. Our data complement these small real-world studies that tofacitinib continues to be efficacious in clinical practice. We found that the response was robust in those who had high disease activity at baseline. In the small number of patients who had low disease activity, the mild increase may represent a regression to the mean. Importantly, the patients who switched from innovator tofacitinib to the generic tofacitinib continued to improve. Although the numbers in this subset are very small to formally evaluate non-inferiority, this is heartening initial data.

We chose a pragmatic definition of failure based on actual events, viz. rheumatologist-prescribed change or drug withdrawal flares: outside of a formal trial, disease activity scores are unlikely to influence clinical decision-making substantially. In addition, many patients were already doing well before starting tofacitinib and were moved over due to financial- or convenience-based (oral versus injectable) reasons, thus making the interpretation of change in disease activity scores more difficult. This line of reasoning was supported by traditional outcome measures: DAS28 worsened in those who were labelled failures; failures were often late and, thus, were important to critically evaluate drug response even after initial response; and, mean survival was longer than that seen in a European registry (112 days).^(15)^ In other studies, disease duration and the number of prior bDMARDs have been associated with tofacitinib non-response.^(12,13,16)^

All adverse events observed were minor. A recent meta-analysis of 20 trials showed a risk ratio of 2.75 for serious infections, higher in the 20 mg/day dose group.^(17)^ TB was the most common opportunistic infection with 26 events noted in a follow-up of more than 5,000 patients; 80% from countries with a high background incidence of TB.^(18)^

TB incidence was 1.21 per 100 per year in Indian patients from clinical trials; interestingly, all those who developed TB were not amongst the 23 patients who had LTBI on screening.^(9)^ Given our substantive number of TB, screening for LTBI may be prudent in this population.

All COVID-19 were mild in these patients. It is tempting to speculate that the tofacitinib may have helped by modulating the inflammatory pathways that produce lung injury and other complications in COVID-19 ^(19)^ These Indian patients also demonstrated the metabolic effects of weight gain^(20)^ and lipid abnormalities^(21)^ seen in other cohorts. Long-term follow-up would help study translation into cardiovascular events, especially when weighed against the increased cardiovascular risk of uncontrolled RA.

To the best of our knowledge, this is the first study reporting the real-world experience of generic tofacitinib from India. Inclusion of patients from a single centre, using tofacitinib monotherapy by the same manufacturer in all reduced possible confounders. Our study has limitations, viz patients were followed up at inconstant time intervals, there is no control group (making attributing causality of both response and AEs difficult). Finally, subgroups (such as patients switching over from innovator tofacitinib) were small, limiting the interpretability.

In summary, generic tofacitinib has shown good effectiveness and tolerable adverse reaction profile despite the high endemicity of infections in India. These findings, make a case for formal long-term real-world studies from India, especially given the widespread use of generic tofacitinib.

## Data Availability

De-identified data will be available with Dr S N Amin for sharing after publication and will be shared depending on the merit and prior approval of proposals. Data requests can be made to

